# SLOTMFound: Foundation-Based Diagnosis of Multiple Sclerosis Using Retinal SLO Imaging and OCT Thickness-maps

**DOI:** 10.1101/2025.07.14.25331522

**Authors:** Reyhaneh Esmailizadeh, Ali Aghababei, Sayeh Mirzaei, Roya Arian, Raheleh Kafieh

**Affiliations:** School of Engineering Science, College of Engineering, University of Tehran, Tehran, Iran; Medical Image and Signal Processing Research Center, Isfahan University of Medical Sciences, Isfahan, Iran; Department of Engineering, Durham University, South Road, Durham, UK

**Keywords:** Foundation Models, Multiple Sclerosis, Optical Coherence Tomography, Scanning Laser Ophthalmoscopy, Artificial Intelligence, Deep Learning

## Abstract

Multiple Sclerosis (MS) is a chronic autoimmune disorder of the central nervous system that can lead to significant neurological disability. Retinal imaging—particularly Scanning Laser Ophthalmoscopy (SLO) and Optical Coherence Tomography (OCT)—provides valuable biomarkers for early MS diagnosis through non-invasive visualization of neurodegenerative changes. This study proposes a foundation-based bi-modal classification framework that integrates SLO images and OCT-derived retinal thickness maps for MS diagnosis. To facilitate this, we introduce two modality-specific foundation models—SLOFound and TMFound—fine-tuned from the RETFound-Fundus backbone using an independent dataset of 203 healthy eyes, acquired at Noor Ophthalmology Hospital with the Heidelberg Spectralis HRA+OCT system. This dataset, which contains only normal cases, was used exclusively for encoder adaptation and is entirely disjoint from the classification dataset. For the classification stage, we use a separate dataset comprising IR-SLO images from 32 MS patients and 70 healthy controls, collected at the Kashani Comprehensive MS Center in Isfahan, Iran. We first assess OCT-derived maps layer-wise and identify the Ganglion Cell–Inner Plexiform Layer (GCIPL) as the most informative for MS detection. All subsequent analyses utilize GCIPL thickness maps in conjunction with SLO images. Experimental evaluations on the MS classification dataset demonstrate that our foundation-based bi-modal model outperforms unimodal variants and a prior ResNet-based state-of-the-art model, achieving a classification accuracy of 97.37%, with perfect sensitivity (100%). These results highlight the effectiveness of leveraging pre-trained foundation models, even when fine-tuned on limited data, to build robust, efficient, and generalizable diagnostic tools for MS in medical imaging contexts where labeled datasets are often scarce.

## 1. INTRODUCTION

Multiple sclerosis (MS) is the most prevalent inflammatory neurological disorder affecting young adults, characterized by demyelination, chronic inflammation, gliosis, and axonal degeneration within the central nervous system (CNS) [1]. Current diagnostic protocols rely predominantly on magnetic resonance imaging and cerebrospinal fluid analysis, which are frequently costly, time-intensive, and inaccessible in resource-limited healthcare environments [2]. Accumulating evidence demonstrates that pathological neurodegenerative changes extend beyond the CNS to encompass the retina of MS patients [3]. Due to shared embryological development, similar tissue architecture, and comparable vascular networks with the brain, the retina represents a direct extension of the CNS, thereby serving as “a window to the brain” [4]. Consequently, retinal imaging modalities, including fundus photography and optical coherence tomography (OCT), present compelling opportunities for non-invasive, cost-effective, and readily accessible MS diagnosis. As Petzold et al. demonstrated in their systematic review [3], OCT investigations have consistently revealed retinal nerve fiber layer (RNFL) and ganglion cell-inner plexiform layer (GCIPL) thinning in MS patients, even in the absence of clinically detectable acute inflammatory episodes (optic neuritis) [3, 4].

The advent of machine learning (ML) has revolutionized medical diagnostics across numerous specialties, with ophthalmology experiencing particularly remarkable advances. ML models have demonstrated diagnostic accuracy levels that rival or exceed those of expert clinicians in detecting and predicting disease progression across a spectrum of ocular pathologies, including diabetic retinopathy, glaucoma, and age-related macular degeneration—the three leading causes of global blindness [5]. While MS primarily manifests as a neurological condition, its association with characteristic retinal pathological changes has prompted investigators to explore ML-based classification approaches using retinal imaging data, yielding promising preliminary results. A comprehensive systematic review and meta-analysis published in 2022 by Nabizadeh et al. showed that ML models trained on OCT data achieve MS detection accuracy levels reaching 93% (sensitivity = 92%, specificity = 93%) [6]. Additionally, infrared reflectance scanning laser ophthalmoscopy (IR-SLO) has recently emerged as a complementary imaging modality, producing comparable diagnostic performance [7, 8]. Aghababaei and colleagues evaluated multiple state-of-the-art convolutional neural network (CNN) architectures, including VGG16, VGG19, ResNet50, and InceptionV3, alongside a convolutional autoencoder combined with simple classification algorithms, on IR-SLO images from both healthy individuals (n = 150) and MS patients (n = 164), achieving optimal classification accuracy of 88% [7]. Extending this work, Arian et al. developed a multimodal approach that integrated IR-SLO images with OCT thickness maps to detect MS using various CNN architectures, including VGG16, VGG19, ResNet50, ResNet101, and a custom CNN model. Their findings demonstrated that the integration of OCT and IR-SLO modalities yielded superior diagnostic performance (accuracy = 92.4 ± 4.1%, area under the receiver operating characteristic curve [AUROC] = 0.97 ± 0.03%) compared to single-modality approaches [8].

Foundation models, a term formally introduced in 2021, encompass artificial intelligence architectures pre-trained on large-scale unlabeled datasets often using self-supervised learning techniques, which can subsequently be fine-tuned for diverse downstream applications [9]. This approach enables models to acquire high-level, generalizable representations without requiring labor-intensive human annotation—a critical advantage in medical domains where vast quantities of imaging data exist but labeled datasets remain scarce due to the expertise needed for accurate annotation [9].

In the ophthalmology domain, Zhou et al. introduced the first retinal foundation model, RETFound, in 2023 [10]. This architecture leverages 1.6 million retinal images, encompassing both OCT scans and fundus photographs, sourced from Moorfields Eye Hospital and University College London. RETFound employs a vision transformer backbone with self-supervised learning through a masked autoencoder [11] strategy: the encoder processes unmasked patches (16×16 pixels) to generate 1024-dimensional feature vectors, while the decoder reconstructs original image patches by integrating these features with masked patch tokens. Zhou et al. showed that the resulting encoder, capable of extracting rich retinal feature representations, can demonstrate remarkable versatility across numerous downstream tasks, including classification of ocular diseases like diabetic retinopathy and glaucoma, AMD progression forecasting, and even detection of systemic conditions such as cardiovascular diseases, stroke, and Parkinson’s disease [10].

Following the development of RETFound., subsequent investigations have been conducted to evaluate its performance across several ophthalmological diagnostic and prognostic applications [12, 13, 14, 15]. For glaucoma detection, Chuter and colleagues validated RETFound using approximately 10,000 fundus photographs from a diverse patient population spanning different ages, ethnicities, and disease stages, achieving an AUROC of 0.86 [12]. Additionally, Chen et al. applied RETFound to fundus photographs (n = 776) to predict two key glaucoma metrics, i.e., cup-to-disc ratio (CDR) and RNFL thickness measurements. Their results were impressive, with R^2^ values of 0.90 for CDR and 0.96 for RNFL thickness predictions. These outcomes also surpassed those of established architectures, including VGG16 (R^2^ = 0.50 for CDR and 0.60 for average RNFL thickness predictions) and vision transformer (R^2^ = 0.69 for CDR and 0.73 for average RNFL thickness predictions) models [14]. In diabetic retinopathy classification, Kuo and colleagues demonstrated RETFound’s notable performance using OCT B-scans from 1,150 participants, achieving 91.2% ± 2.3% accuracy. This represented a satisfactory improvement over ResNet50 (82.5% ± 5.1%) and vision transformer (84.9% ± 2.3%) architectures [13]. Zhang et al. conducted a comprehensive community screening study in Shanghai, China, analyzing over 17,000 fundus photographs from healthy individuals and patients with AMD, diabetic retinopathy, and pathologic myopia. RETFound achieved high sensitivity rates of 95% for diabetic retinopathy detection, 76% for AMD, and 100% for pathologic myopia, outperforming both EfficientNetB3 and ResNet50 models [15]. Collectively, these studies establish RETFound as a transformative tool in ocular disease classification and prognostic evaluation, demonstrating clear superiority over conventional CNN models utilizing supervised learning paradigms.

Building upon prior studies on IR-SLO and OCT integration, which demonstrated that combining SLO images and OCT thickness maps enhances the accuracy of MS diagnosis [8], we propose a novel bi-modal framework leveraging these modalities in conjunction with foundation models. Notably, this study addresses several important gaps in current knowledge. First, existing artificial intelligence systems for MS classification using retinal markers have relied exclusively on traditional ML and CNN approaches, overlooking the revolutionary capabilities of foundation models. This research represents the pioneering application of a foundation model to retinal imaging for MS detection. Second, previous studies utilizing RETFound have employed either fundus photographs or OCT B-scans as standalone imaging modalities, without exploring their combined potential. This investigation addresses this gap by examining whether the integration of IR-SLO—which shares visual characteristics with fundus photographs—and OCT can improve MS diagnostic performance using RETFound, beyond what each modality achieves individually. Third, this work uniquely applies RETFound to OCT thickness maps rather than conventional B-scans. While ocular diseases such as diabetic retinopathy, AMD, or glaucoma produce structural changes readily observable in B-scan cross-sections, neurodegenerative conditions like MS primarily manifest as quantitative reductions in retinal layer thickness, particularly affecting the RNFL and GCIPL. Therefore, the utilization of OCT thickness maps represents a more appropriate and targeted approach for detecting MS-related retinal changes.

The remainder of this paper is organized as follows: Section 2 introduces the proposed foundation-based bi-modal classification framework, detailing the architecture, modality integration, and fine-tuning strategies. Section 3 outlines the experimental setup, presents the evaluation results. Sections 4 and 5 provide a discussion and a conclusion of the findings, respectively, and emphasize the advantages of leveraging foundation models. Finally, Section 6 presents details regarding code availability.

## 2. MATERIALS AND METHODS

### 2.1 Preprocessing

To leverage the complementary strengths of both SLO images and OCT thickness maps for each eye, we begin by extracting these modalities and applying a series of preprocessing steps to prepare them as suitable inputs for the proposed bi-modal model. The SLO images are readily available for each eye; however, generating the corresponding OCT thickness maps requires a dedicated processing pipeline, illustrated in Figure 1. This figure illustrates the process of generating OCT thickness maps for each eye. For visualization, only five b-scans are shown, although the actual dataset includes a significantly larger number.

**Figure 1.**
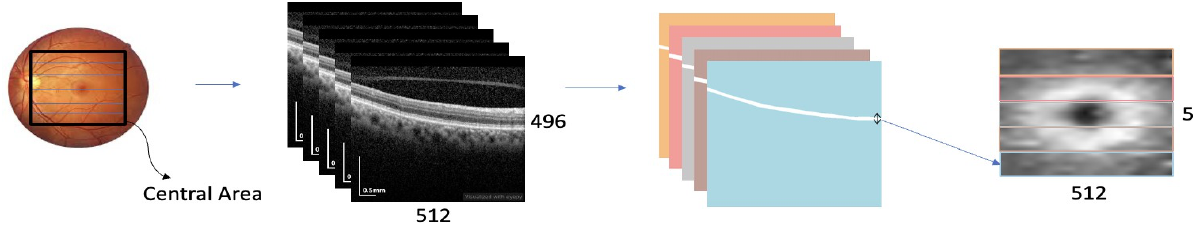
Processing pipeline for generating OCT thickness maps.

The thickness maps are generated by first identifying the retinal layer of interest within the OCT volume. For each lateral position, the thickness is calculated by subtracting the coordinate of the upper boundary of the specified retinal layer from that of the lower boundary in each B-scan. These per-slice thickness values, computed across all B-scans, are then aggregated to form a 2D thickness map that captures the spatial distribution of the selected retinal layer over the scan area.

In this study, we focused on evaluating which retinal layers provide the most relevant structural information for MS diagnosis using OCT thickness maps. Based on prior evidence, the ganglion cell–inner plexiform layer (GCIPL), retinal nerve fiber layer (RNFL), and whole retinal thickness have each been associated with neurodegenerative changes [3, 4]. To assess their relative diagnostic value, we generated thickness maps for each of these layers and evaluated their impact on classification performance within the OCT configuration. This targeted analysis allowed us to identify the most informative retinal layer(s) for MS detection within our framework. All resulting thickness maps were resized to a standardized spatial resolution of 224 × 224 pixels with three channels and subsequently normalized using modality-specific mean and standard deviation values. Similarly, all SLO images were resized and normalized following the same procedure to ensure consistency across input modalities in the bi-modal pipeline.

### 2.2 Custom Foundation Models for SLO Images and Thickness Maps

In this study, we aim to harness the capabilities of foundation models to advance the diagnosis of multiple sclerosis (MS) through retinal imaging analysis. RETFound [10], a state-of-the-art vision transformer–based foundation model specifically developed for retinal imaging, serves as the backbone of our proposed framework. It has been pre-trained on an extensive dataset of over 1.6 million retinal images using a self-supervised learning strategy grounded in the Masked Autoencoders (MAE) paradigm. This large-scale pretraining enables RETFound to capture rich, hierarchical representations of retinal anatomy and pathology, which are essential for robust feature extraction.

Two pretrained variants of the RETFound model are publicly available: one trained on the ImageNet-1K dataset and OCT B-scans (RETFound-OCT) and the other on the ImageNet-1K dataset and color fundus photographs (RETFound-Fundus). Given that both SLO images and OCT-derived thickness maps share greater visual and structural similarity with fundus images than with raw OCT B-scans, we opted to build upon the RETFound-Fundus model for our work. However, since RETFound-Fundus was originally trained exclusively on fundus images, we performed modality-specific fine-tuning to adapt the model for our input data. Specifically, we fine-tuned RETFound-Fundus separately on SLO images and thickness maps to better align the learned feature representations with each modality. As a result, we obtained two customized foundation models optimized for these respective image types. Importantly, this targeted fine-tuning approach—leveraging a relatively limited amount of domain-specific data—allowed us to repurpose a powerful foundation model without the need for pretraining from scratch on millions of images. This significantly reduces computational demands while enabling effective adaptation to new modalities.

#### 2.2.1 Data Collection for New Foundation Models

To develop modality-specific foundation models, we fine-tuned the RETFound-Fundus model using an independent dataset that, while entirely distinct from our primary classification dataset and acquired from a different clinical center, shares a comparable distribution in terms of imaging characteristics. However, the fine-tuning dataset consists exclusively of normal eyes. It comprises 203 eyes from healthy Iranian individuals and was acquired using the Heidelberg Spectralis HRA+OCT system (Spectralis HRA+OCT; Heidelberg Engineering, Heidelberg, Germany) at Noor Ophthalmology Hospital [16]. Although the dataset used in this stage is fully labeled, we employed a self-supervised approach to generate new foundation models, using no labels during this phase.

#### 2.2.2 SLOFound: Self-Supervised Pretraining on SLO Images

As depicted in Figure 2 and previously described, the RETFound-Fundus was pretrained on ImageNet-1K and further adapted using large-scale color fundus images. SLOFound was then obtained by fine-tuning RETFound-Fundus on SLO images from our dataset of the Noor Ophthalmology Hospital. SLOFound adopts a masked image modeling approach using a masked autoencoder (MAE) architecture. During training, random patches of each SLO image are masked, and the model learns to reconstruct the missing regions from the visible context. This self-supervised learning strategy allows the model to capture robust and transferable retinal features specific to the SLO modality, without requiring manual annotations.

**Figure 2.**
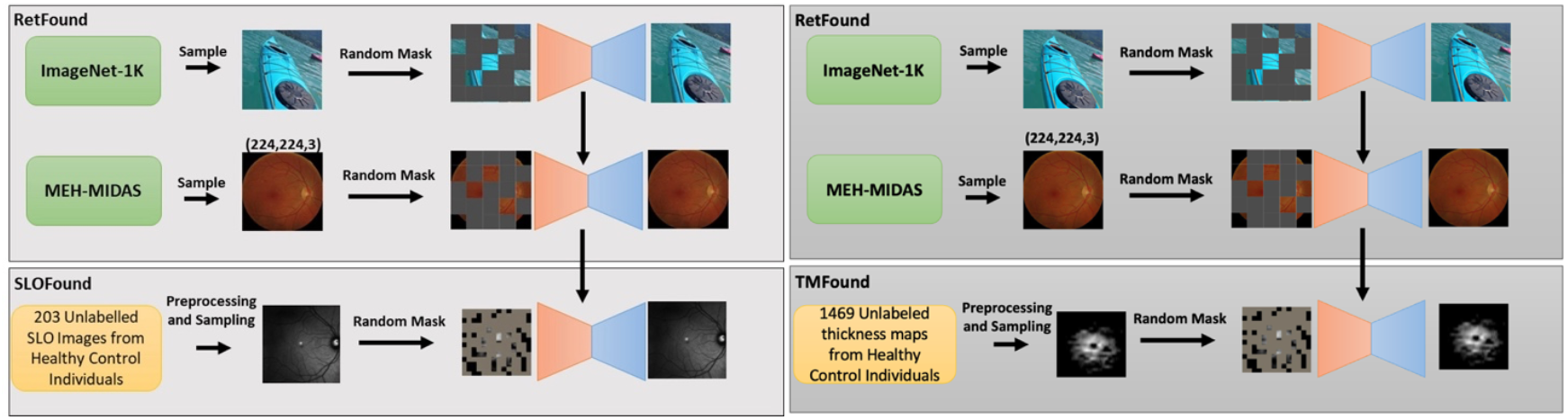
SLOFound and TMFound generation.

#### 2.2.3 TMFound: Self-Supervised Pretraining on Thickness Maps

The development of the TMFound model followed the same masked autoencoding-based self-supervised learning framework described for SLOFound. However, in this case, the input modality consisted of retinal thickness maps derived according to the methodology described in the preprocessing section. 1,469 thickness maps of different retina layers were extracted from 203 healthy eyes. The complete training pipeline for TMFound is depicted in Figure 2.

### 2.3 Foundation-Based Bi-Modal Framework

With the SLOFound and TMFound models trained, we utilize their respective encoders to independently extract high-level semantic features from SLO images and retinal thickness maps. These features encapsulate modality-specific information that is critical for fine-grained classification. To harness the complementary strengths of both modalities, we implement a late fusion strategy: each encoder outputs a 1024-dimensional feature vector, and the resulting vectors are concatenated to form a unified representation. This fused feature vector is subsequently passed to a downstream classifier for final MS prediction. The proposed bimodal integration enhances overall diagnostic performance and improves generalization in MS detection tasks.

#### 2.3.1 Classification Dataset

For the classification task, we used IR-SLO images from 32 patients with MS and 70 healthy controls (HC), acquired with a SPECTRALIS SD-OCT device (Heidelberg Engineering, Germany), as part of a previous study by Ashtari et al. (2017– 2019) at the Kashani Comprehensive MS Center in Isfahan, Iran [17]. OCT B-scans were evaluated using Heidelberg Eye Explorer (HEYEX v5.1) and met OSCAR-IB quality standards [18]. The automatic real-time mean (ART) algorithm averaged nine repeated scans per location to reduce speckle and background noise. Retinal layers were segmented using a graph-based texture method and manually corrected by an expert. Each 6 × 6 mm macular volume consisted of 45 horizontal B-scans with 512 A-scans per line and 3.8 µm axial resolution, stored in .vol format.

To assess model generalization, and consistent with our previous study, 20% of the subjects from the Isfahan dataset were reserved as an independent test set. The remaining data were used for training and validation with an 80/20 split ratio. To prevent data leakage and ensure robust evaluation, a subject-wise splitting strategy was adopted, ensuring that all images from the same individual, regardless of eye laterality, were assigned exclusively to either the independent testing set or the remaining data.

#### 2.3.2 Classifier Architecture and Tuning

The architecture of the fully connected component of the bi-modal classifier, including the number of hidden layers and their corresponding neurons, was optimized through hyperparameter tuning using the Optuna framework over 20 trials [19]. Optimization was performed on pairs of labeled SLO images and their corresponding retinal thickness maps. The overall structure of the proposed bi-modal model is illustrated in Figure 3.

**Figure 3.**
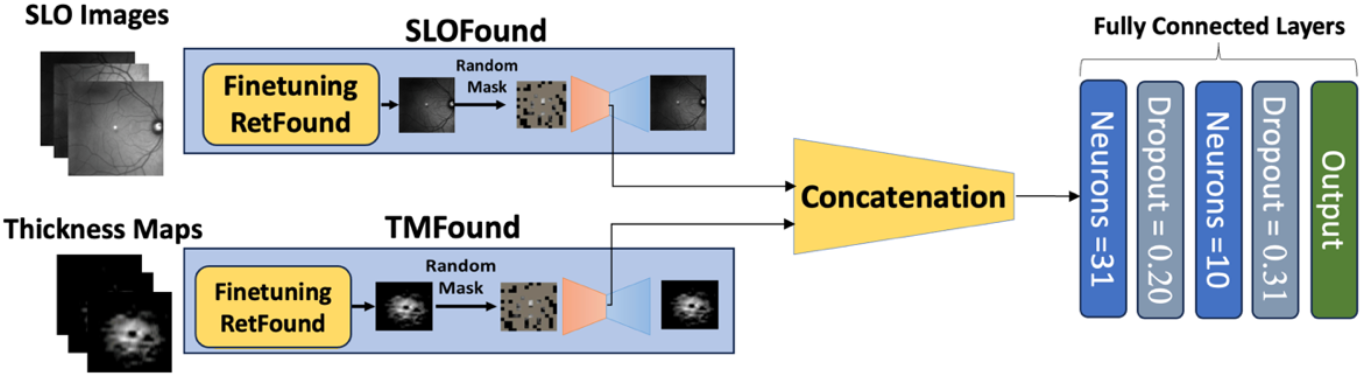
Overall architecture of the proposed bi-modal classification model.

#### 2.3.3 Evaluation Metrics

The performance of the proposed model was evaluated using several standard classification metrics, including accuracy (ACC), sensitivity (SE), specificity (SP), precision (PR), and F1-score (F1). The mathematical definitions of these metrics are provided below, where TP, FN, TN, and FP denote true positives, false negatives, true negatives, and false positives, respectively.

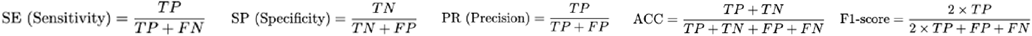

In addition, the area under the ROC curve (AUROC) was computed as a summary measure of the model’s overall discriminative ability.

## 3. RESULTS

All experiments in this study were conducted in a cloud-based environment equipped with an NVIDIA Tesla T4 GPU and 16 GB of RAM. The training and evaluation pipelines were implemented in Python using the PyTorch framework, and hyperparameter tuning was performed using Optuna.

### 3.1 Identifying the Most Informative Retinal Layer for MS Diagnosis

To identify the most informative retinal layer for MS diagnosis, we evaluated the performance of the TMFound (unimodal) model using thickness maps derived from three individual layers: the ganglion cell–inner plexiform layer (GCIPL), the retinal nerve fiber layer (RNFL), and the whole retina. In addition, we assessed a combined configuration incorporating all three layers.

In our training strategy, the TMFound encoder was unfrozen and fine-tuned using a layer-wise learning rate decay schedule. In this approach, layers closer to the output were assigned higher learning rates, allowing them to adapt more aggressively during training, while earlier layers received progressively smaller updates. This reflects the principle that lower layers capture general features and should remain relatively stable, whereas higher layers are more task-specific and benefit from greater flexibility during fine-tuning. The decay rate across layers was set to 0.6. This hierarchical optimization strategy facilitates the effective adaptation of pretrained representations to the downstream classification task. The classification results for each configuration are presented in Table 1, allowing for a direct comparison of their diagnostic value.

**Table 1.** Performance Metrics Across Retinal Layers and Their Combination.

| Metrics | GCIPL | RNFL | Whole Retina | GCIPL+RNFL+Retina |
| --- | --- | --- | --- | --- |
| **ACC** | **0.9211** | 0.8158 | 0.8158 | 0.8158 |
| **AUC-roc** | **0.9970** | 0.9851 | 0.9851 | 0.9940 |
| **F1-Score** | **0.9183** | 0.8146 | 0.8146 | 0.8146 |
| **Sensitivity** | **0.9375** | 0.8542 | 0.8542 | 0.8542 |
| **Specificity** | **0.9375** | 0.8542 | 0.8542 | 0.8542 |
| **Precision** | **0.9118** | 0.8333 | 0.8333 | 0.8333 |

As evident from Table 1, the GCIPL layer consistently outperforms other retinal layers across all evaluation metrics. Therefore, we focus on this layer for the remainder of the manuscript.

### 3.2 Performance of the Proposed Foundational-Based Bi-Modal Classifier

In this stage, we incorporated SLO images to take advantage of the complementary information provided by this modality alongside the features used in the previous stage. The strategy involved freezing the weights of the SLOFound and TMFound. Additionally, the parameters of the classifier were initialized using Glorot initialization (also known as Xavier initialization) [20], which helps maintain consistent variance of activations across layers and promotes stable and efficient training dynamics.

The result obtained from the proposed foundation-based bi-modal model is highly promising, demonstrating superior performance across all key evaluation metrics. The bi-modal model consistently outperforms the uni-modal (TMFound-based) configuration, underscoring the benefits of integrating complementary modalities for more accurate and reliable MS classification. The loss and accuracy curves for validation and training datasets are shown in Figure 4.

**Figure 4.**
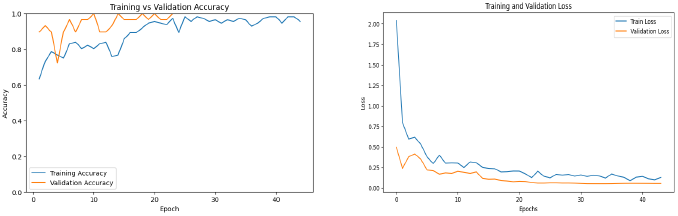
Loss and accuracy curves for the proposed foundation-based bi-modal model.

### 3.3 Comparison with Prior State-of-the-Art Resnet-Based Model

We compare the performance of the proposed foundation-based bi-modal model with our previously developed ResNet-based model, which established the state-of-the-art for MS classification on this dataset [8]. Both models utilize the same two modalities—SLO images and retinal thickness maps—but differ significantly in architectural design. The earlier approach employed ResNet-based encoders for feature extraction, whereas the current model leverages pretrained foundation model encoders (SLOFound and TMFound). This architectural advancement promotes more effective feature representation and improved generalization.

As shown in Table 2, the proposed foundation-based bi-modal model slightly outperforms the previous ResNet-based model in accuracy (0.9737 vs. 0.9685) and specificity (0.9583 vs. 0.9496), while maintaining perfect sensitivity (1.0). Although the ResNet-based model has a slightly higher AUC-ROC and precision, the superior overall accuracy and balanced performance of the proposed model highlight its robustness. Accuracy provides a balanced summary of classification performance across both classes and is particularly important in binary medical diagnosis tasks, where both false positives and false negatives carry clinical consequences.

**Table 2.** Performance comparison between the previous ResNet-based bi-modal classifier and the proposed foundation-based bi-modal model.

| Metric | ResNet-Based Model | Proposed Foundation-Based Model |
| --- | --- | --- |
| Accuracy (ACC) | 0.9685 | **0.9737** |
| AUC-ROC | **0.9969** | 0.9792 |
| F1-Score | **0.9683** | 0.9655 |
| Sensitivity (Recall) | 1.0000 | 1.0000 |
| Specificity | 0.9496 | **0.9583** |
| Precision | **0.9522** | 0.9333 |

### 3.4 Model Interpretability

Model interpretability for images refers to the ability to understand and explain how a machine learning model, particularly a deep neural network, arrives at its predictions when processing visual data. It involves techniques that reveal which parts of an image contribute most to a model’s decision, helping researchers and practitioners assess the model’s reasoning, trustworthiness, and potential biases. To support this, a variety of interpretability techniques have been developed. Among them is the Transformer Explainability algorithm [21], which introduces a novel method by assigning local relevance using the Deep Taylor Decomposition framework and propagating these scores across the model’s layers. By applying the Transformer Explainability algorithm to the best-performing unimodal model trained with GCIPL thickness maps, the saliency maps visualized in Figure 5 were generated. For each subject in this Figure, the left image shows the GCIPL thickness map, the middle image shows the MS class activation map, and the right image shows the HC class activation map. The top two predicted classes, corresponding logits (value), and class probabilities predicted by the best-performing unimodal model are displayed above each subject.

**Figure 5.**
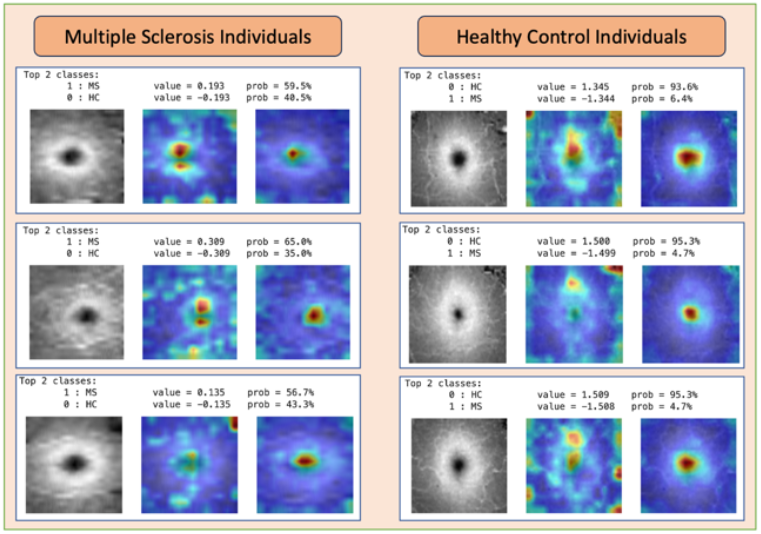
Visualization results for 3 individuals with Multiple Sclerosis (MS) and 3 Healthy Controls (HC) from the test set.

## 4. DISCUSSION

This study had several limitations. First, the classification dataset was relatively small and obtained from a single center, potentially limiting the model’s generalizability. To address this, we introduced SLOFound and TMFound—foundation models known for their robustness and reliability when fine-tuned on limited data. Moreover, the dataset used for self-supervised training of the masked autoencoders that generated SLOFound and TMFound consisted solely of healthy control (HC) subjects, raising concerns about the models’ ability to capture features representative of multiple sclerosis (MS). We mitigated this limitation during downstream classification by incorporating a separate dataset comprising 32 MS patients out of 102 total subjects.

## 5. CONCLUSION

In this study, we introduced two foundation models—SLOFound and TMFound —designed specifically for Scanning Laser Ophthalmoscopy (SLO) images and Optical Coherence Tomography (OCT)-derived retinal thickness maps, respectively. These models were adapted by fine-tuning the RetFound backbone on a relatively small dataset, thus reducing the extensive data requirements typically associated with training foundation models from scratch. The proposed bi-modal classification framework integrates these pre-trained encoders to enable effective fine-tuning. To inform model design, we first systematically assessed the diagnostic value of various retinal layers and identified the GCIPL as the most informative for MS classification. Based on this finding, all subsequent development and evaluation leveraged GCIPL-derived thickness maps in combination with SLO images. Experimental results demonstrate that our foundation-based bi-modal model surpasses both unimodal configurations and a prior ResNet-based state-of-the-art model, particularly in terms of classification accuracy and specificity. These improvements highlight the strength of foundation model adaptation, as both SLOFound and TMFound were fine-tuned from RetFound using a relatively small dataset, yet delivered superior performance compared to traditionally trained architectures. This underscores the promise of leveraging pre-trained foundation models in medical imaging domains where annotated data is limited, enabling more efficient, accurate, and generalizable diagnostic tools for MS.

## Data Availability

All data produced in the present study are available upon reasonable request to the corresponding authors

https://github.com/Reyhaneesmailizadeh/SLOTMFound

## 6. CODE AVAILABILITY

Our article code and weights for SLOFound and TMFound are available here.

## Notes

### Competing Interest Statement

The authors have declared no competing interest.

### Funding Statement

This study did not receive any funding

### Author Declarations

Ethics committee/IRB of National Institute for Medical Research and Development (NIMAD) gave ethical approval for this work.

### Summary of Updates

Footer added to the first page to clarify the status of the paper. Page number added to each page.

## REFERENCES

[1] M.T. Wallin et al. “Global, regional, and national burden of multiple sclerosis 1990–2016: a systematic analysis for the Global Burden of Disease Study 2016”. In: The Lancet Neurology 18 (2019), pp. 269–285. DOI: 10.1016/S1474-4422(18)30443-5.

[2] A.J. Thompson et al. “Diagnosis of multiple sclerosis: 2017 revisions of the McDonald criteria”. In: Lancet Neurology 17 (2018), pp. 162–173. DOI: 10.1016/S1474-4422(17)30470-2.

[3] A. Petzold et al. “Retinal layer segmentation in multiple sclerosis: a systematic review and meta-analysis”. In: Lancet Neurology 16 (2017), pp. 797–812. DOI: 10.1016/S1474-4422(17)30278-8.

[4] J. Lambe, S. Saidha, and R.A. Bermel. “Optical coherence tomography and multiple sclerosis: Update on clinical application and role in clinical trials”. In: Multiple Sclerosis 26 (2020), pp. 624–639. DOI: 10.1177/1352458519872751.

[5] D.S.W. Ting et al. “Artificial intelligence and deep learning in ophthalmology”. In: British Journal of Ophthal-mology 103 (2019), pp. 167–175. DOI: 10.1136/bjophthalmol-2018-313173.

[6] F. Nabizadeh et al. “Diagnostic performance of artificial intelligence in multiple sclerosis: a systematic review and meta-analysis”. In: Neurological Sciences (2022). DOI: 10.1007/s10072-022-06460-7.

[7] A. Aghababaei et al. “Discrimination of multiple sclerosis using scanning laser ophthalmoscopy images with autoencoder-based feature extraction”. In: Multiple Sclerosis and Related Disorders 88 (2024), p. 105743. DOI: 10.1016/j.msard.2024.105743.

[8] Roya Arian et al. “SLO-Net: Enhancing Multiple Sclerosis Diagnosis Beyond Optical Coherence Tomogra-phy Using Infrared Reflectance Scanning Laser Ophthalmoscopy Images”. In: Translational Vision Science & Technology 13.7 (2024), p. 13. DOI: 10.1167/tvst.13.7.13.

[9] R. Bommasani et al. On the Opportunities and Risks of Foundation Models. arXiv preprint. 2022. DOI: 10.48550/arXiv.2108.07258. eprint: arXiv:2108.07258.

[10] Yukun Zhou et al. “A foundation model for generalizable disease detection from retinal images”. In: Nature 622.7981 (2023), pp. 156–163. DOI: 10.1038/s41586-023-06682-7.

[11] K. He et al. Masked Autoencoders Are Scalable Vision Learners. arXiv preprint. 2021. DOI: 10.48550/arXiv.2111.06377. eprint: arXiv:2111.06377.

[12] B. Chuter et al. “Evaluating a Foundation Artificial Intelligence Model for Glaucoma Detection Using Color Fundus Photographs”. In: Ophthalmology Science 5 (2025), p. 100623. DOI: 10.1016/j.xops.2024.100623.

[13] D. Kuo et al. “How Foundational Is the Retina Foundation Model? Estimating RETFound’s Label Efficiency on Binary Classification of Normal versus Abnormal OCT Images”. In: Ophthalmology Science 5 (2025), p. 100707. DOI: 10.1016/j.xops.2025.100707.

[14] J. Zhang et al. “RETFound-enhanced community-based fundus disease screening: real-world evidence and decision curve analysis”. In: NPJ Digital Medicine 7 (2024), p. 108. DOI: 10.1038/s41746-024-01109-5.

[15] M.S. Chen et al. “Independent Evaluation of RETFound Foundation Model’s Performance on Optic Nerve Analysis Using Fundus Photography”. In: Ophthalmology Science 5 (2025), p. 100720. DOI: 10.1016/j.xops.2025.100720.

[16] Raheleh Kafieh et al. “Thickness Mapping of Eleven Retinal Layers Segmented Using the Diffusion Maps Method in Normal Eyes”. In: Journal of Ophthalmology 2015 (2015), pp. 1–11. DOI: 10.1155/2015/259123.

[17] Fereshteh Ashtari, Ali Ataei, Raheleh Kafieh, et al. “Optical coherence tomography in neuromyelitis optica spectrum disorder and multiple sclerosis: a population-based study”. In: Multiple Sclerosis and Related Disorders 47 (2021), p. 102625. DOI: 10.1016/j.msard.2021.102625.

[18] S. Schippling, L. Balk, F. Costello, et al. “Quality control for retinal OCT in multiple sclerosis: validation of the OSCAR-IB criteria”. In: Multiple Sclerosis Journal 21.2 (2015), pp. 163–170. DOI: 10.1177/1352458514538110.

[19] Takuya Akiba et al. Optuna: A Next-Generation Hyperparameter Optimization Framework. 2019. DOI: 10.48550/arXiv.1907.10902. arXiv: 1907.10902 [cs.LG]. URL: https://arxiv.org/abs/1907.10902.

[20] Xavier Glorot and Yoshua Bengio. “Understanding the difficulty of training deep feedforward neural networks”. In: Proceedings of the Thirteenth International Conference on Artificial Intelligence and Statistics (AISTATS). Vol. 9. 2010, pp. 249–256.

[21] Hila Chefer, Shir Gur, and Lior Wolf. “Transformer Interpretability Beyond Attention Visualization”. In: Pro-ceedings of the IEEE/CVF Conference on Computer Vision and Pattern Recognition (CVPR). June 2021, pp. 782–791.

